# Quantifying the rate and magnitude of the Omicron outbreak in China after sudden exit from ‘zero-COVID’ restrictions

**DOI:** 10.1101/2023.02.10.23285776

**Authors:** Emma E. Goldberg, Qianying Lin, Ethan O. Romero-Severson, Ruian Ke

## Abstract

In late 2022, China transitioned from a strict ‘zero-COVID’ policy to rapidly abandoning nearly all interventions and data reporting. This raised great concern about the presumably-rapid but undisclosed spread of the SARS-CoV-2 Omicron variant in a very large population of very low pre-existing immunity. A quantitative understanding of the epidemic dynamics of COVID-19 during this period is urgently needed. Here, by modeling a combination of case count and survey data, we show that Omicron spread extremely fast, at a rate of 0.42/day (95% CrI: [0.35, 0.51]/day) after the full exit from zero-COVID policies on Dec. 7, 2022. Consequently, we estimate that the vast majority of the population (97%, 95% CrI [95%, 99%]) was infected during December, with the nation-wide epidemic peaking on Dec. 23. Overall, our results highlight the extremely high transmissibility of the variant and the importance of proper design of intervention exit strategies to avoid large infection waves.

## Introduction

From the beginning of the SARS-CoV-2 pandemic until fall 2022, China maintained a strict ‘zero-COVID’ set of policies such as frequent testing and large scale lockdowns that kept the number of infected individuals at very low levels. Beginning on Nov. 11, 2022, these control measures were rapidly relaxed, culminating in nearly all intervention efforts being abandoned after Dec. 7. This abrupt exit from zero-COVID raised public health concerns about the unchecked spread of SARS-CoV-2 [1], especially given the high transmissibility of the Omicron variants BA.5 and BF.7 present in China [2, 3]. Indeed, a few weeks after the full exit from zero-COVID, it was reported that high numbers of patients with respiratory illness were overwhelming hospitals in China [4]. Furthermore, on three flights from China to Italy in late Dec. 2022, ∼40%–50% of passengers tested positive for COVID-19 [5, 6]. These observations strongly suggest that SARS-CoV-2 was already widespread in China by the end of Dec. 2022, in contrast to the official data that showed daily cases during December to be low and waning (Fig. 1A). However, even with these anecdotal reports, we still do not have formal estimates of the rate and the magnitude of the outbreak in China.

**Figure 1:**
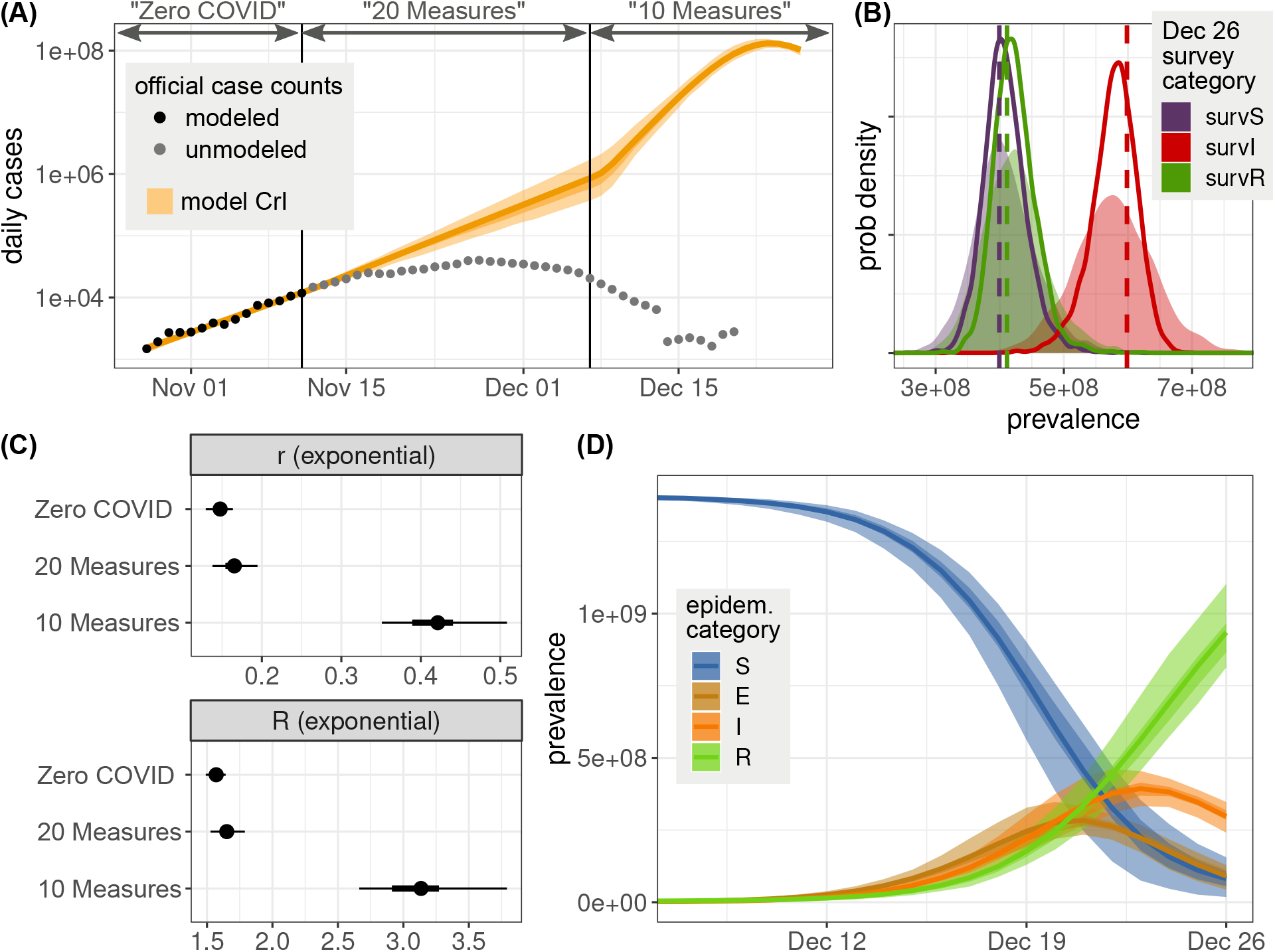
Model-predicted epidemic dynamics of the Omicron variant in China. **(A)** Official case counts compared with the model-inferred true number of cases near the end of 2022. Shades of orange show the median, 50% CrI, and 95% CrI. **(B)** Survey results, interpreted as population size, compared against model-predicted values for those survey categories on Dec. 26. Dashed lines are the data, solid curves are the posterior distribution of expected number of individuals, and shaded curves are the posterior predicted number of individuals. **(C)** Estimates of the intrinsic rate of increase, *r*, and reproductive number, *R* during the exponential growth of each time period. Points are the median, thick lines are the 95% CrI, and thin lines are the 50% CrI. (See also Table S1.) **(D)** Model-inferred epidemic dynamics, during 20 Measures. The median, 50% CrI, and 95% CrI are shown for people in the model’s susceptible (S), exposed (E), infected (I), and recovered (R) states (cf. Fig. S1).

Accurately quantifying the dynamics of the Omicron outbreak in China is urgently needed for several reasons. First, it is essential to understand the magnitude of the public health crisis induced by large-scale SARS-CoV-2 spread in a population of size greater than 1.4 billion. Second, it will reveal the efficacy of the previously-strict non-pharmaceutical intervention efforts during late Oct. and early Nov. in China. This in turn can help to design effective combinations of pharmaceutical and non-pharmaceutical intervention policies to better ‘flatten the curve’ of future waves [1, 7]. And finally, the population in China likely had little immune protection against the Omicron variant, as we show below. The epidemic in China provided a unique situation for directly estimating the ‘intrinsic’ transmissibility of the Omicron variant, which has been a central issue in understanding its evolution [8, 9, 10].

The major difficulty in understanding the Omicron epidemic in China is the lack of data directly tracking the spread. Compulsory mass testing and reporting were gradually stopped from Nov. 11 onward, and as we show below, the official numbers of confirmed cases subsequently do not reflect the true extent of SARS-CoV-2 spread. Indirect data must therefore be used. Recent modeling work used subway ridership and online infection status survey data to estimate the dynamics of the SARS-CoV-2 epidemic in Beijing [11]. However, its conclusions of most rapid SARS-CoV-2 spread around Nov. 17 (well before full exit from zero-COVID) and dual epidemic peaks in mid-Dec. are at odds with the changes in policies and later data released by China Center for Disease Control and Prevention (China CDC) [3]. Furthermore, it is unknown whether the predicted epidemic trajectories in Beijing can be generalized to understand the epidemic throughout the country. Clearly, analysis of more extensive data are still needed to understand the outbreak in China as a whole.

Here, we quantify the Omicron epidemic dynamics in China by drawing on results from online surveys of COVID-19 infection status conducted by a website for the Chinese health authorities on Dec. 26 [12]. Despite its limitations (discussed below), the survey represents the currently-best evidence of the extent of SARS-CoV-2 spread in China by this time. We developed a modeling approach that integrates both the official case count data before Nov. 11 and the Dec. 26 survey data to reveal a more complete picture of the dynamics of SARS-CoV-2 before and after the dismantling of zero-COVID. Extensive sensitivity analyses confirm that our conclusions are robust to many assumptions about the data and model, and our findings are validated by comparison against two other independent datasets.

## Results

### Understanding the official case count data during the different phases of public health policy

We first aimed to understand the spread of the Omicron variant in China immediately before the relaxation of zero-COVID (i.e., between Oct. 28 and Nov. 11, 2022). Under zero-COVID, a wide variety of intervention efforts were implemented stringently, including frequent city-wide lock-downs, travel and movement restrictions, and compulsory quarantine of infected individuals and their contacts in centralized facilities. During this period before Nov. 11, testing was mandatory and frequent, leading the official case count data to be a reliable portrayal of the levels of COVID-19 infection in China. Despite the strict measures, with the extremely transmissible Omicron variants, the data show a clear exponential increase in COVID-19 cases shortly before the relaxation of zero-COVID (Fig. 1A). Using a negative binomial regression (see Methods), we estimated the rate of this exponential growth to be 0.14/day (95% CI: [0.13, 0.15]/day) during this period.

Official data after Nov. 11 report that growth of the SARS-CoV-2 epidemic slowed (Fig. 1A). For example, using only the official case counts we estimated the rate of exponential growth between Nov. 12 and 26 to be 0.06/day (95% CI: [0.05, 0.07/day), substantially lower than the growth rate estimated during zero-COVID. Peaking on Nov. 27, the official daily reported cases declined to levels consistent with the zero-COVID policy by early December. This pattern is unexpected because after Nov. 11, the government started to relax some interventions, implementing the ‘20 measures’ policy from Nov. 12 to Dec. 6 and the ‘10 measures’ policy on Dec. 7, leading to a full exit from the previous zero-COVID policy. Presumably, the relaxation of such strict interventions would lead to more rapid spread, contrary to what is observed from the official case count data. We reason that the only plausible explanation for the lower and lower growth rates after Nov. 11 is the cessation of the requirement for mass testing and rigorous reporting, which led to a rapid decoupling of the surveillance data from the epidemic intensity. As a result, the official case count data after Nov. 11 reflect the rapidly declining detection rate rather than real change in the infection dynamics.

### Estimating SARS-CoV-2 transmission dynamics by integrating multiple sources of data

To account for the under-reported official case count data after Nov. 11, we collected results from an online survey conducted by a website for the Chinese Ministry of Human Resources and Social Security on Dec. 26, 2022 (see Methods for details). In this online survey, 47,897 participants across China reported whether they had tested positive or thought they were currently infected with SARS-CoV-2, and whether they had COVID-19 symptoms or had recovered from the symptoms on the day of survey. The survey thus estimated the fraction of individuals in three categories: those who reported they had never been infected, those who reported that they currently had COVID-19 symptoms, and those who reported they had been but were no longer symptomatic. Note that these survey categories do not directly correspond to the epidemiological states of susceptible, infected, and recovered, because each person is reporting based on symptoms and does not know their true infectiousness status. When interpreted correctly, however, these data should provide insight into the epidemic stage and thus allow inference of the epidemic dynamics.

We developed an expanded susceptible-exposed-infected-recovered (SEIR) model to include states directly corresponding to categories in the data that we used for fitting and for validation (Fig. S1; see Methods for details). Separating the model into three time periods (Fig. 1) corresponding to changes in official policy (late Oct. to Nov. 11: zero-COVID; Nov. 12 to Dec. 7: 20 Measures; and Dec. 8 onward: 10 Measures) allowed us to estimate temporal changes in the transmission rates. We assumed a constant transmission parameter within each time period for simplicity. We fit to the official case counts, which we believe to be reliable up to Nov. 11, to estimate the transmission rate for the first period. In the second period we do not have a clear observation to estimate the transmission rate directly, so we simply assumed that it must be equal to or greater than the rate in the first period due to the ongoing relaxation of intervention policies. The dynamics of the third period is then mainly determined by the population survey data on Dec. 26. Other model parameters, such as the incubation period and duration of symptoms, were assumed the same for all time periods and were fixed to values obtained from the literature (see Methods).

Overall, we estimated that the outbreak in China grew exponentially at rates of 0.15/day (95% CrI [0.13, 0.15] /day) under zero-COVID between Oct. 28 and Nov. 11 (Fig. 1C). This translates to an epidemic doubling time of 4.7 days (95% CrI [4.5, 5.4] days). Under the 20 Measures policy, the epidemic grew at 0.17/day (95% CrI [0.14, 0.17] /day), translating an epidemic doubling time of 4.2 days (95% CrI [4.0, 5.0] days). After the full exit from zero-COVID, the outbreak initially grew exponentially at a rate of 0.42/day (95% CrI [0.35, 0.44] /day), and the epidemic doubling time was 1.6 days (95% CrI [1.6, 2.0] days). Then the epidemic peaked around Dec. 23 (Fig. 1D). Taking the generation interval distribution and parameters from the literature [13], we calculated the reproductive number, *R*, during the exponential increase periods under and after the full exit from zero-COVID (using Eq. (2) in Methods) as 1.57 (95% CrI [1.49, 1.59]) and 3.13 (95% CrI [2.66, 3.27]), respectively (Fig. 1C). Based on these values, we estimated that the zero-COVID policies prior to relaxation suppressed the transmission of these Omicron variants by 56% (95% CrI [0.46, 0.64]).

Our results suggest that on Dec. 7, i.e. the day when the ‘10 Measures’ policy (full exit from zero-COVID) was announced, there were already approximately 1 million new infections (Fig. 1C). Because of the extremely high rate of spread (with a doubling time of 1.6 days) afterwards, the outbreak increased to very large sizes later in Dec. Indeed, our model estimated that 97% (95% CrI [95%, 99%]) of the population (i.e. 1.37 billion people) became infected in December, and as a result of the exponential nature of the spread, the vast majority of people (88% of the population, 95% CrI [83%, 93%]) became infected during the short window of time between Dec. 15 and 31, 2022 (Fig. 1D).

### Robustness to model and data assumptions

Our main analysis necessarily makes many assumptions about the epidemiological processes and the case count and survey data. We tested whether our main conclusions hold under variants of those assumptions, particularly looking to test our finding that nearly the entire population became infected in Dec. 2022.

First, we previously assumed that the entire population of China was fully susceptible to Omicron infection. To test the validity of this assumption, we calculated the level of population immunity against Omicron in 2022, based on published vaccination data [14] and characteristics of vaccine effectiveness against infection [15] (see Methods). We estimated that the population immunity is likely to be below 0.1% (Fig. S3). Our assumption of complete immunity is therefore essentially valid.

Second, we previously assumed that the numbers of confirmed cases from Oct. 28 to Nov. 11 represented the actual number of infected individuals, i.e., that the case reporting rate in China was 100%. This is a reasonable approximation because of the mandatory mass testing in place: all individuals were required to get tested every 2–5 days. However, it is possible that some infected individuals were not detected. We therefore considered an alternative in which the reporting rate was only 50%. Here, the estimated growth rate after Dec. 7 was reduced slightly to 0.403/day, and the epidemic again peaked on Dec. 23.

Third, we previously assumed the Dec. 26 survey results were an unbiased random sample of the entire population. If, however, recovered people were more likely to respond to the survey, or people reported symptoms not due to COVID-19 (e.g., instead due to influenza - an unlikely scenario given the recent report by China CDC [3]), the numbers of symptomatic or recovered would be over-reported. We therefore made a large perturbation of the Dec. 26 data by moving 20% of people from each of the symptomatic and recovered categories into the uninfected category. The estimated growth rate was then reduced somewhat to 0.39/day after Dec. 7, the size of the susceptible population by Dec. 31 remained below 4%, and the epidemic peak moved slightly to Dec. 24.

Fourth, we previously assumed that the entire population is well-mixed. If, however, people who live in areas with high transmission were also more likely to be represented in the data, our results could over-state the total number of infected people by failing to recognize subpopulations that remain uninfected. For example, people who live in dense urban areas with good internet access and health care facilities may be more likely to be included in COVID-19 testing and online survey, while also experiencing more opportunity for infection. We therefore added to our model a second set of states representing susceptible, exposed, infected, and recovered people in a subpopulation that is not sampled. This subpopulation was assigned a lower transmission rate (half that of the sampled population), and transmission between the two subpopulations was assumed to be even lower (one tenth of the transmission rate). We partitioned the entire country’s population to be 65% sampled and 35% unsampled, reflecting the nationwide urban-rural proportion. Our results from this model were also entirely consistent with our main findings, with an epidemic peak on Dec. 23 and only 2.6% of the entire population remaining susceptible by the end of Dec.

Fifth, we took many parameter values from the literature. We therefore tested the sensitivity of our results to those assumed values by re-fitting the model using different values. Even when setting the value of each state transition parameter (*k*_*E*_, *k*_*IP*_, *k*_*IS*_, *k*_*S*_, *k*_*A*_, and *k*_*T*_ ; all defined in Fig. S1) to either 25% higher or lower than our baseline value, the median exponential growth rate remained between 0.38–0.50 /day after Dec. 7, the size of the population still susceptible by Dec. 31 remained less than 4%, and the epidemic peak remained between Dec. 21–24 (Fig. S4).

### Model validation using additional datasets

The estimation above for the periods after Nov. 11 is heavily dependent on the Dec. 26 prevalence survey data. We therefore sought to validate our results by comparing them against datasets that were not used in the inference above.

First, as we were preparing this manuscript, the China CDC released a set of data on voluntary PCR and antigen tests and the test positivity rate across local hospital networks in China for most of Dec. 2022 and Jan. 2023 [3]. These data are likely not a representative sample of the infection status of the population as a whole, since people who suspect they might be infected are probably more likely to engage in voluntary testing. But although these data may not directly reveal the magnitude of cases, they can reveal the pace and the peak timing of the epidemic. To compare our model predictions against this new dataset, we adjusted our predictions to reflect the nature of voluntary tests. Specifically, we multiplied the model-predicted case counts (representing the true, complete incidence) by an approximation of the testing effort and the fact that recovered individuals are less likely to get tested again (see Methods and Eq. (4)). We found surprisingly close agreement between our adjusted model predictions and the national testing data in Dec. 2022 (Fig. 2).

**Figure 2:**
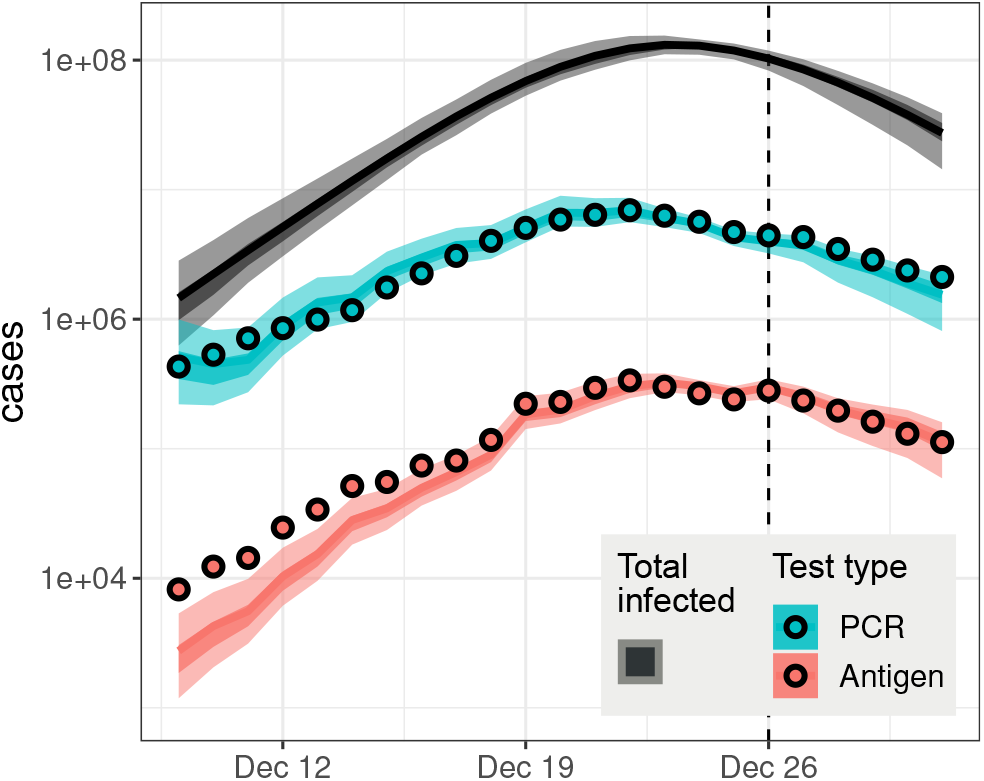
Model comparison with the national testing survey by China CDC. The gray band shows the complete incidence inferred by our main model fit, which did not include the data points plotted here. Those model predictions adjusted for testing effort and willingness to test are shown by colored bands, for PCR and antigen tests. The adjustment procedure involved one free parameter for each test type; details are provided in Methods. For the model predictions, shading shows the median, 50% CrI, and 95% CrI.

Second, we collected data from a large survey conducted by the Sichuan CDC. Survey participants in Sichuan province were asked about their infection status, the date of test positivity, and the date of symptom onset. We decided these data could not be incorporated into our main model fit because the epidemic in Sichuan was ahead of much of the country, as judged by this province containing the fourth-highest proportion of recovered individuals in the Dec. 26 national survey (Table S2). However, we compared the trend of the spread in this dataset with our estimates. We estimated the exponential growth rates of the epidemic in Sichuan in early Dec. by the date of test positivity or the date of symptom onset to be 0.37/day or 0.43/day, respectively (Fig. S5). These estimated growth rates are remarkably consistent with our estimates above for the country as a whole.

Overall, our model results based on two data sources—official case counts and the Dec. 26 survey—agree very well with two additional data sources, strongly supporting the reliability of our conclusions.

## Discussion

We modeled the epidemic dynamics of the Omicron variant of SARS-CoV-2 in China from Nov. to Dec. 2022, during a period when China moved from having the the world’s strictest COVID-19 policies (‘zero-COVID’), to little-to-no intervention efforts. We found that in the period directly preceding the end of zero-COVID, the Omicron epidemic was already growing exponentially at a rate of 0.15/day. After full exit from zero-COVID, the Omicron variant spread at a very high rate of 0.42/day, with a doubling time of 1.6 days, during early and mid-Dec. before the incidence peaked around Dec. 23. This rate of spread is much quicker than the rate of spread of the original Wuhan strain, which we previously estimated to be 0.29/day (doubling time of 2.4 days) [16].

Because of the extremely high rate of exponential spread, we estimated that the vast majority of people in China (approximately 90% of the population, i.e., greater than 1 billion people) became infected during a short period of time between Dec. 15 and 31, 2022. This would lead to a large number of people needing health care, far exceeding the hospital capacity, and thus explains the report that hospitals were overwhelmed during this period [4]. Furthermore, we estimated that at least 95% of the population (1.3 billion people) was infected during December. With an infection fatality ratio between 0.1–0.2% for the Omicron variant [17], we would expect between 1.3 to 2.6 million COVID-19 deaths in China during Dec. 2022 as well as Jan. 2023 (because of the delay from infection to death). These results emphasize the need for gradually relaxing intervention efforts (instead of abruptly changing policy) and implementing additional measures (e.g., pharmaceutical strategies) to ‘flatten the curve’ [7]. This would help to ease hospital burden, ensure sufficient health care for infected people, prevent epidemic overshoot (i.e. a large final epidemic size because of rapid spread [18]), and ultimately reduce the number of deaths.

Our results differ from a recent paper by Leung et al. [11] that modeled the epidemic dynamics in Beijing over a similar period. First, they estimated that the highest rate of epidemic growth occurred in mid-Nov., a period when substantial intervention efforts were still in place, whereas our estimates suggest the rate of epidemic growth was highest after the full exit from zero-COVID. The model used by Leung et al. [11] assumes that the overall contact rate is a function of the daily number of subway travelers, which peaked mid-November. However, it is unclear how the number of subway passengers is related to transmission, given that a large fraction of transmission likely occurs in the household [19]. More generally, it has been shown that human mobility after the first wave of the epidemic in 2020 relates to changes in transmission in complex ways [20]; the relationship between the mobility measures and transmission can be insignificant in many countries, country-specific, or intervention policy-specific. As a result, the results in Leung et al. [11] could be biased by this strong assumption. Second, Leung et al. [11] estimated that the epidemic incidence in Beijing had two peaks (one on Dec. 10 and another on Dec. 21); however, we estimated the epidemic in China peaked on Dec. 23, remarkably consistent with the newly-released data from China CDC [3] (Fig. 2).

One substantial limitation of our analysis is that it is heavily dependent on the online infection status survey data on Dec. 26. Although it is conducted by an official source, there could be many factors biasing the results. We directly addressed two concerns about these data. First, the survey could over-state the progression of the epidemic if people with symptoms are more likely to take the survey, or if people mistake influenza for COVID-19 symptoms. Recent data from the China CDC report [3] shows that among individuals who had influenza-like illnesses and were tested for both COVID-19 and influenza, only a very small fraction were positive for influenza, especially in late Dec. Nevertheless, we artifically increased the proportion of uninfected people in the survey and obtained very similar results. Second, because the survey was conducted online, the demographic characteristics of the participants may be different from the general population. In particular, if a substantial subpopulation did not participate in the survey and also experienced lower rates of spread—for example, people living in sparsely-populated areas—our estimate of the total number of people infected could be too high. We tested this assumption by fitting a structured population model and obtained very similar results. Indeed, the data released by China CDC [3] show that across different provinces, indicators of COVID-19 infection, including positive tests and fever clinic visits, decreased continuously throughout Jan. 2023, strongly suggesting that the fraction of susceptible individuals was at a very low level throughout the country by the end of Dec., as predicted by our model. Finally, we validated our model findings against other data sources that were not used in our model inference. We found strong concordance with both the estimated peak time of the epidemic and its growth trajectory. These many lines of evidence strongly suggest that our estimation is reliable and robust, despite the limitations of the available data.

The rate of spread we estimated for the period after full exit from zero-COVID is higher than the rate of Omicron spread estimated in many other countries, for example, in England [21]. This is at least partially because, unlike most other countries where there existed sizable levels of population immunity against the Omicron variant arising from recent natural infection and/or vaccination, the population in China had very little to no protection against the Omicron variant. The vaccinations delivered in China were based on the original Wuhan strain and were typically administered 8 months prior to Dec. 2022 [14]. Because of low levels of antibody titres against Omicron and their rapid decline after vaccination [15], our analysis shows that immune protection against this variant was likely negligible in the population. Therefore, the growth rate and the reproductive number (0.42/day and 3.13, respectively) during the exponential growth period we estimated may be a good approximation of the intrinsic transmissibility of the variants BA.5 and BF.7 in a densely populated area without population immunity. When we compare these estimates to the estimates for the original strain during the outbreak in Wuhan, China in Jan. 2020 (0.29/day and 5.7, respectively) [16], the results indicate that the Omicron variant evolved a higher intrinsic transmission fitness by shortening the time needed for transmission to occur (and thus the generation interval), rather than generating more secondary infections (and obtaining a high reproductive number).

Quantifying the growth characteristics before and after zero-COVID, we estimated these intervention efforts reduced the transmission of the Omicron variant by approximately 56%. As a comparison, the effect of lock-down during the spread of the original variant in early 2020 was estimated to be 70–80% in Europe [22]. Interestingly, the Omicron outbreak in China had already been growing in Nov. 2022 before the relaxation of zero-COVID (albeit at a low rate). This may reflect the high transmissibilty of the variant and that the stringent measures of the zero-COVID policy were not effectively implemented because of factors such as population noncompliance due to pandemic fatigue. Nonetheless, these results highlight the difficulty of containing a respiratory infection that causes explosive outbreaks and transmits during the presymptomatic or asymptomatic period.

Because the epidemic grew so rapidly in Dec., we may expect another sizable wave of infection later in 2023 when the large cohort of people infected at the end of 2022 become susceptible to reinfection. The timing and magnitude of the future wave would depend on how quickly the population immunity wanes over time [23, 24, 25] and the ability of newly emerging variants to evade the immunity caused by the Omicron variants [26, 27]. Therefore, modeling efforts that evaluate intervention strategies, including vaccination, will be crucial to reduce the level of infection and mortality.

## Methods

### Datasets and sources

#### Official case counts

National Health Commission (NHC) of the People’s Republic of China had been releasing reports on COVID-19 infections from the beginning of the pandemic until Dec. 23, 2022 [28]. These reports include numbers of symptomatic, asymptomatic, imported, recovered infections, and deaths across the country. Because the definitions of ‘symptomatic’ and ‘asymptomatic’ infections have been changing over time, we use only the total number of cases by summing the two categories. We retrieved all official data from Sina Pandemic Map [29], which provides pre-processed and well-organized daily case data from NHC. Note the NHC data is also used as a source for the Johns Hopkins COVID-19 Dashboard [30].

#### Prevalence survey on Dec. 26, 2022

On Dec. 26, 2022, the RenSheTong online platform (https://m12333.cn), a website for the Chinese Ministry of Human Resources and Social Security, conducted a one-day online survey of the COVID-19 infection status for people living in China [12]. There were 47,897 participants in total. In the questionnaire, participants were asked about their infection status and their duration of symptoms. Infection status was reported in four categories. ‘Uninfected’ was defined as individuals who had never tested positive or were unaware they were infected. ‘Asymptomatic’ was defined as individuals who had tested positive, but did not have symptoms of infection at the time of the survey. ‘Symptomatic’ was defined as individuals who had tested positive or thought they were infected, and were experiencing symptoms at the time of the survey. ‘Recovered’ was defined as individuals who had tested positive or thought they had been infected by COVID-19 in the past, and had recovered from symptoms of infection. Data from the survey are reproduced as Table S2. Because of the small number of respondents in the Asymptomatic category, our analyses folded them in with the Susceptible category, which already included individuals who could be infected but without symptoms.

Participants in the survey were also asked about the duration of symptoms. We fitted an Erlang distribution to the survey data by estimating the mean of the distribution and adjusting the shape parameter of the Erlang distribution from 1 to 10.

#### Chinese CDC reports of number of positive tests after Dec. 9, 2022

The Chinese Center for Disease Control and Prevention disclosed national COVID-19 pandemic data on Jan. 25, 2023, including PCR test results, antigen test results, outpatient and inpatient data, variant surveillance data, and vaccination progress across the country [3]. Data were collected from official surveillance systems, labs, hospitals, and mobile applications where residents could upload their antigen test results voluntarily. We used daily numbers of positive PCR and antigen tests and daily test positivity rates.

#### Sichuan survey on Dec. 25 and 26, 2022

The Sichuan Center for Disease Control and Prevention Health conducted an online survey of the COVID-19 infection status for people living in Sichuan province on Dec. 24 and 25 [31]. There were a total of 158,506 participants. In the questionnaire, participants were asked about their infection status and their duration of symptoms. Infection status was categorized as ‘uninfected’ or ‘tested positive either by PCR or antigen test.’ In addition, participants reported the date that they tested positive and the date of their symptom onset.

### Regression to estimate exponential growth rates

We estimated the rate of exponential growth for different time periods nation-wide by fitting a generalized negative binomial regression to the official case count data. Fitting was performed with the glm.nb() function in the R programming environment [32].

The rates of exponential growth in Sichuan province during Dec. 2022 were estimated by fitting a linear regression model to the fractions of COVID-19 positive individuals or individuals who started symptoms between Dec. 2 and Dec. 9. The Dec. 1 data point was ignored because it is likely to include individuals who tested positive or started to have symptoms before Dec. 1. The fitting was performed using the lm() function in the R programming environment [32].

### The expanded SEIR model

We developed an SEIR-type model to encompass not only the daily case counts from extensive official testing, but also the survey data in which respondents’ answers were based on symptoms. In this model, we categorized the population into 12 classes, plus 2 additional classes for bookkeeping: *S* for susceptible, *E*_1_ and *E*_2_ for exposed, *I*_*P*_ for pre-symptomatic and infectious, *I*_*S*_ for symptomatic and infectious, *R*_*S*1_, *R*_*S*2_, and *R*_*S*3_ for symptomatic but no longer infectious, *R*_*S*_ for recovered and no longer with symptoms, *I*_*A*1_ and *I*_*A*2_ for asymptomatic (throughout infection), and *R*_*A*_ for recovered and never having symptoms. States *T*_1_ and *T*_2_ are used to record individuals who test positive, in order to compare with daily case counts. Fig. S1 shows how individuals transit from one class to another. Parameters *k*_*i*_ govern the rates of flow from one class to the next, *f* is the fraction of infected individuals who eventually develop symptoms, and *w* is the proportion of cases found by testing. The system of equations defining this model is:

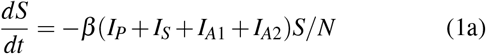

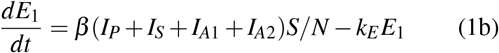

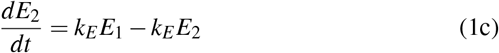

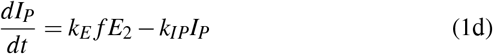

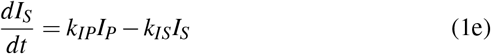

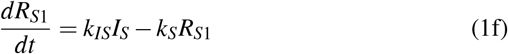

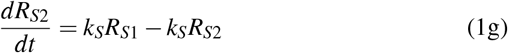

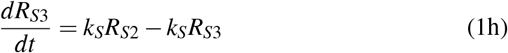

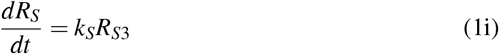

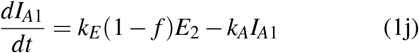

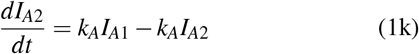

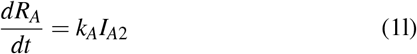

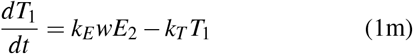

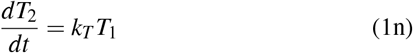

Our model structure allowed us to map our model states to the categories in the Dec. 26 online survey. People who reported themselves ‘uninfected’ could be truly susceptible (*S*), asymptomatic but exposed (*E*_1_, *E*_2_) or infected (*I*_*A*1_, *I*_*A*2_), or recovered never having had symptoms (*R*_*A*_). People who reported themselves ‘infected’ were experiencing symptoms, so they could be still infectious (*I*_*S*_) or no longer infectious (*R*_*S*1_, *R*_*S*2_, *R*_*S*3_). People reporting themselves ‘recovered’ were no longer experiencing symptoms (*R*_*S*_).

### Parameter values and statistical inference

We fixed most of the parameters in the model to values estimated from epidemiological studies. We used an incubation period for Omicron of mean 3.4 days [33] and a generation interval for Omicron of mean 3.3 days [13]. Both distributions can be approximated by a gamma distribution with a shape parameter of 3, so in our model there are three states describing individuals who are infected but yet to develop symptoms (*E*_1_, *E*_2_, and *I*_*P*_). We then set the rate parameters *k*_*E*_ = 2*/*1.9 /day and *k*_*IP*_ = 1*/*1.5 /day so that the mean pre-symptomatic infectious period is 1.5 days [34] and total incubation period is 3.4 days [33], and *k*_*IS*_ = 1*/*1.5 /day so that the mean infectious period after symptom onset is 1.5 days.

We estimated from the population survey data (Table S3) that the distribution of the symptomatic period can be approximated by a gamma distribution with a mean of 5.7 days and a shape parameter of 4 [12]. Therefore, in our model, we used four states to represent the symptomatic period (*I*_*S*_, *R*_*S*1_, *R*_*S*2_ and *R*_*S*3_), and we set *k*_*S*_ = 3*/*4.2 /day such that the mean duration that an individual is in the *R*_*S*1_, *R*_*S*2_ or *R*_*S*3_ state is 4.2 days and thus the mean symptomatic period is 4.2 + 1.5 = 5.7 days.

Our model contained six free parameters: the transmission rate during each of the three periods (*β*_*i*_), the fraction of infected individuals who eventually develop symptoms (*f*), an initial number of exposed individuals in mid-Oct. (*E*_0_), and an overdispersion parameter for the count data (*ϕ*), which were assumed to follow a negative binomial distribution. We used an initial condition for the ODE system of zero prevalence for all states except *E*_1_ = *E*_2_ = *E*_0_ on Oct. 22, one week before the beginning of the case count data we fit. The model was fit using Bayesian inference and implemented in the Stan programming language [35]. We set the prior distribution of each *β*_*i*_ to be normal with mean 3 and standard deviation 2, the prior on *f* to be uniform between 0 and 1, the prior on *E*_0_ to be uniform between 1 and ten times the number of cases on the first day of data, and the prior on *ϕ*^−1^ to be exponential with mean 5. These are weak priors, and the posterior estimates (shown in Fig. S2) were not sensitive to them.

The growth rate, *r*, during the exponential growth period of the outbreak was calculated as the dominant eigenvalue of the Jacobian matrix of the ODE system, Eq. (1), assuming the population is fully susceptible (i.e., *S* = *N*).

### Calculation of the reproductive number, *R*, ***during exponential growth***

We used the growth rates, *r*, of SARS-CoV-2 in China estimated in this study and the distribution of the intrinsic generation interval to calculate *R*. According to Abbott et al. [13], the distribution of the generation interval has a mean of 3.3 days and a shape parameter close to 3. *R* is then calculated using the formula provided in Park et al. [36]:

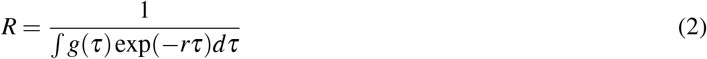

where *g* is the density function of the intrinsic generation interval distribution.

### Estimating population immunity against Omicron infection

Lau et al. [15] estimated vaccine effectiveness (VE) of the CoronaVac vaccine against Omicron infection: 7 days following immunization, the VE for the second and third doses were 5% (0–27%) and 30% (1–66%), respectively, and then the VE reduced exponentially to 1% (0–11%) and 6% (0–29%), respectively, 100 days after immunization. We used the VE function and parameter values derived by Lau et al. [15] in our calculation assuming that all the vaccines administered in China follow the same VE characteristics as CoronaVac. We then collected data on the fractions of people who already received two doses or three doses of vaccine over time from Ref. [14] (Fig. S3A). From this dataset, we estimated the fractions of people newly vaccinated with their second or third dose over the entire period of consideration. Note that the fourth dose of vaccine was not authorized in China until mid-Dec. 2022, and therefore we did not include the fourth dose in our calculation.

Then, we calculated the population immunity against Omicron on day *d, P*(*d*), as

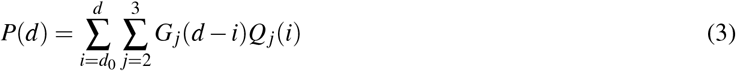

where *Q*_*j*_(*i*) is the fraction newly vaccinated with the second or the third dose (*j* = 2 or 3, respectively) on day *i*, and *G*_*j*_(*d − i*) is the VE on day *d* given an individual received the second or the third dose (*j* = 2 or 3, respectively) on day *i*.

The expected and upper bound estimates (Fig. S3B) were made using the expected and upper bound estimates of the VE.

### Model adjustment for testing intensity and willingness in the China CDC data

We compared the fitted model trajectory against the China CDC data to validate our model estimations. For this, we multiplied the total cases predicted by the model by a factor to correct for testing effort and the willingness to get tested. We defined

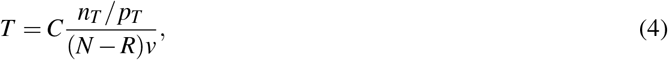

where *T* is the model-adjusted inferred number of positive tests (plotted as colored bands in Fig. 2), *C* is the total number of model-predicted cases (gray band in Fig. 2), *n*_*T*_ is the number of positive tests, *p*_*T*_ is the test positivity rate, *N* is the total country population size, *R* is the number of individuals in the recovered non-symptomatic model states (*R*_*S*_ + *R*_*A*_), and *v* is an unknown factor related to the proportion of the population that engages in voluntary testing. The term (*N − R*)*v* reflects that not all individuals engage in the testing and among those who engage, individuals who know they are recovered likely do not get tested or self-test any more. Values of all the quantities in Eq. (4) are known for each day—from the model fit or the reported data—except for *v*. We estimated a single value of *v* (across all days) for each testing type by least-squares fitting, using Eq. (4), obtaining *v* = 0.44 for the PCR tests and *v* = 0.52 for the antigen tests.

## Data Availability

All data produced in the present study are available upon reasonable request to the authors.

