## Supplementary Figures for "Quantifying the rate and magnitude of the Omicron outbreak in China after sudden exit from ‘zero-COVID’ restrictions"

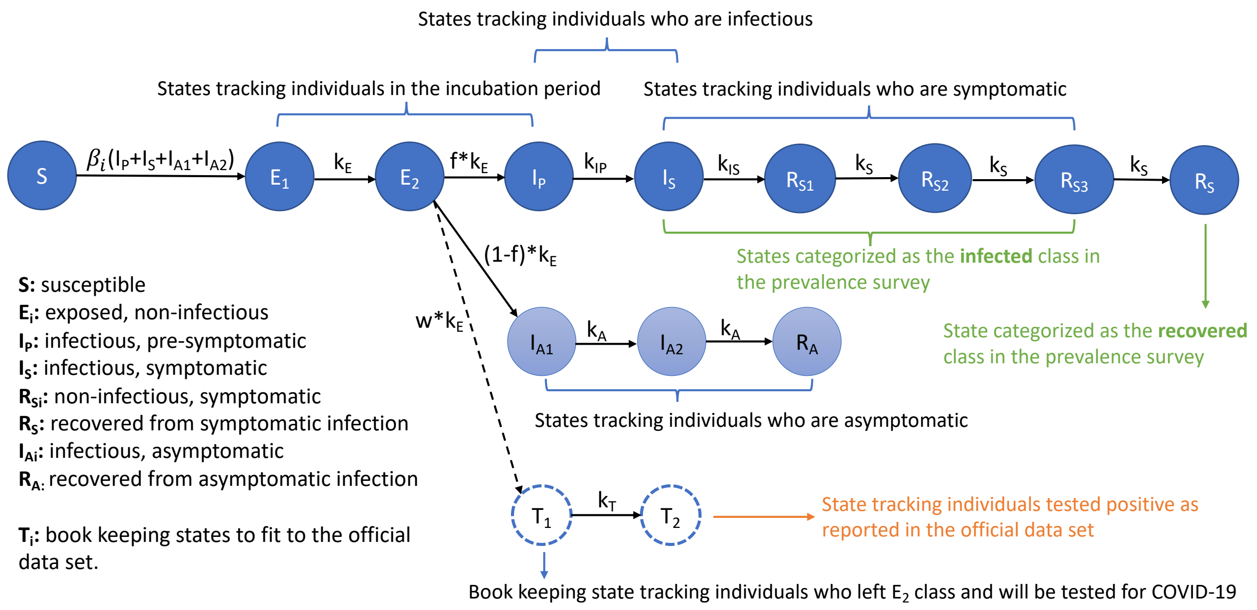


Figure S1: **Schematic and descriptions of the SEIR-type model.** This model is designed to integrate multiple data sources. States that correspond to categories of the datasets were marked with colored texts.

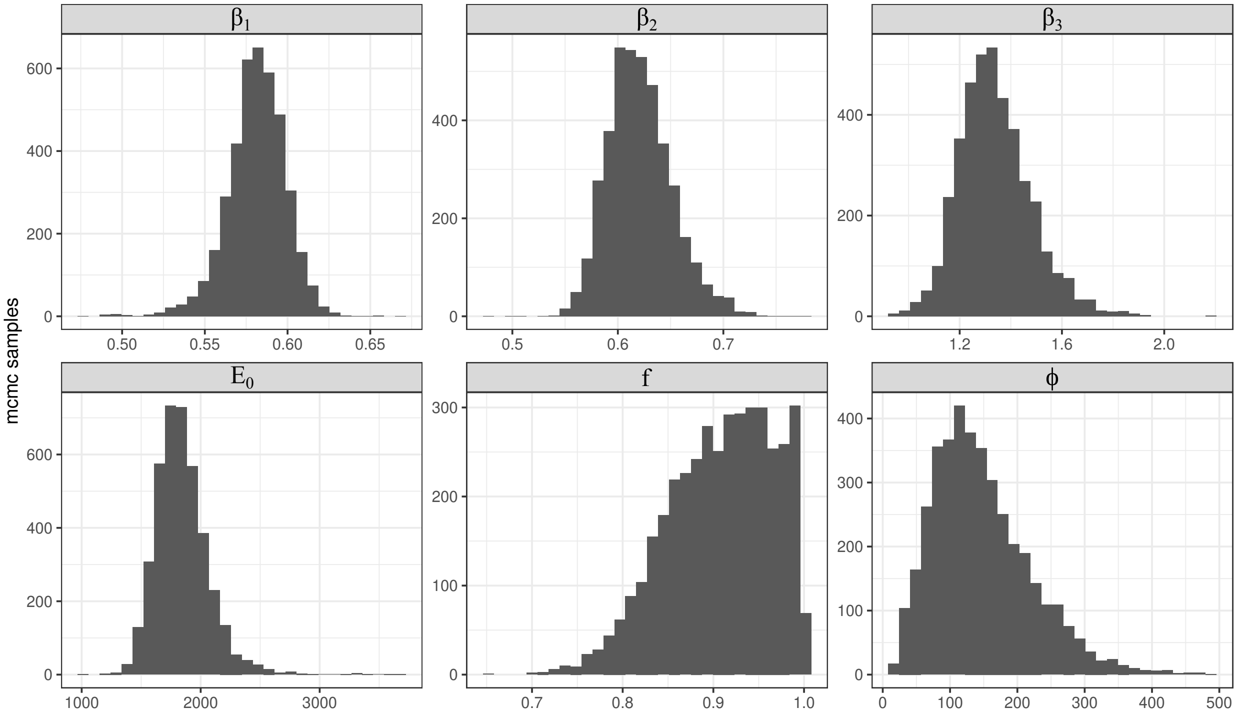


Figure S2: **Posterior distributions of model parameters.** The free parameters in our model were the transmission rate for each time period ($\beta_i$), an initial condition on the number of exposed people on Oct. 22 ($E_0$), the proportion of infected people who eventually develop symptoms ($f$), and an overdispersion parameter for the count data ($\phi$).

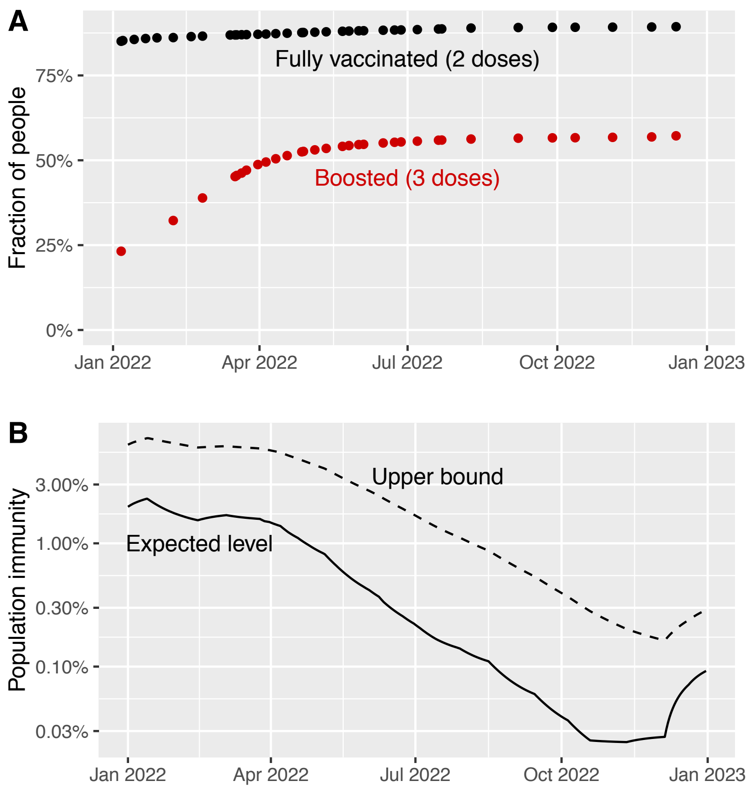


Figure S3: **Population immunity induced by vaccination in China was very low against Omicron infection.** Most individuals received their second or third dose of vaccine prior to 2022 or Apr. 2022, respectively. Combined with relatively low vaccine effectiveness, this led to overall low population immunity, despite efforts to increase vaccination coverage in late 2022. **(A)** Fraction of people vaccinated according to the number of doses received, as reported in Ref. [14]. **(B)** Estimated levels of population immunity against Omicron infection in China using data in (A) and the vaccine effectiveness function in Ref. [15]. Expected values and upper bound values were shown as solid and dashed lines, repsectively. Note that the vertical axis is plotted on a log scale.

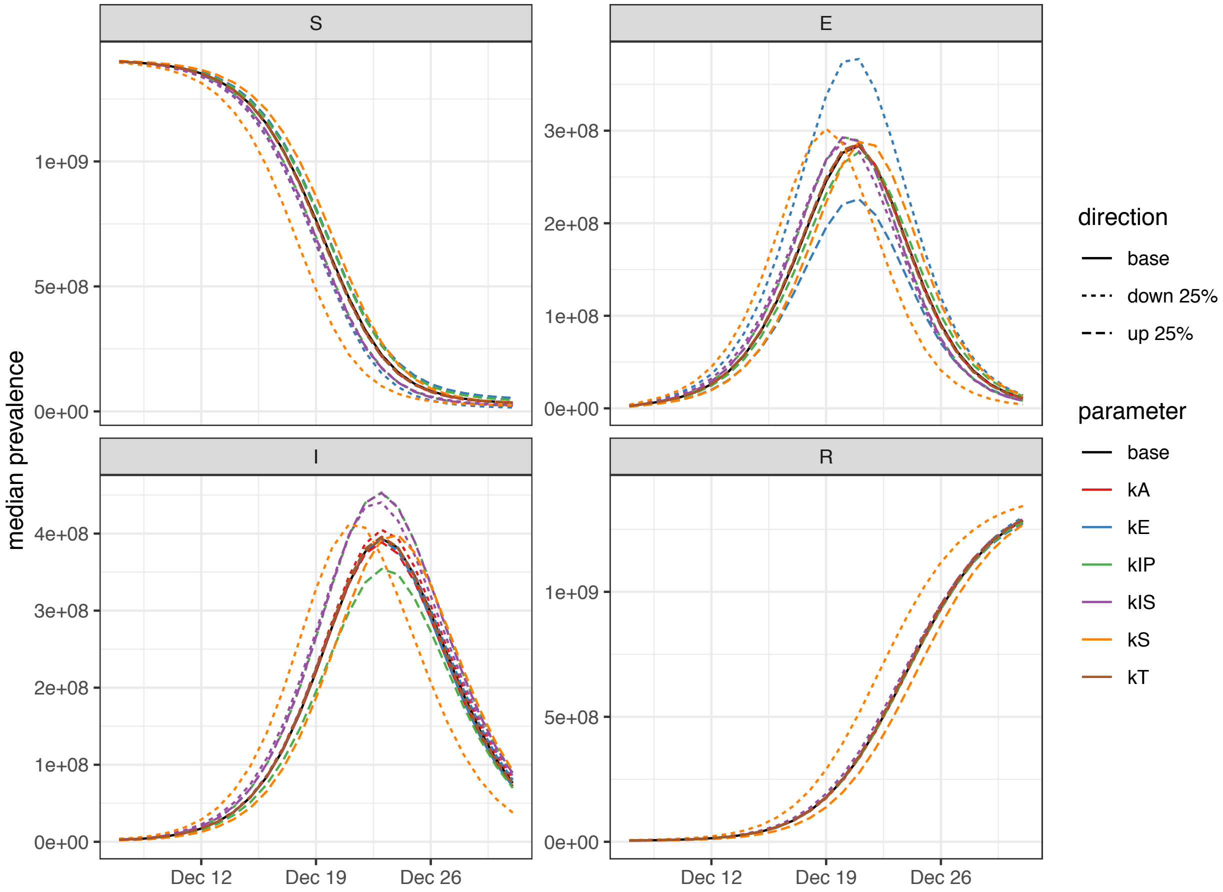


Figure S4: **Sensitivity of inferred epidemiological dynamics to perturbations of the parameter values.** Model parameters are defined in Fig. S1 and Eq. (1), and baseline parameter values are stated in the Methods.

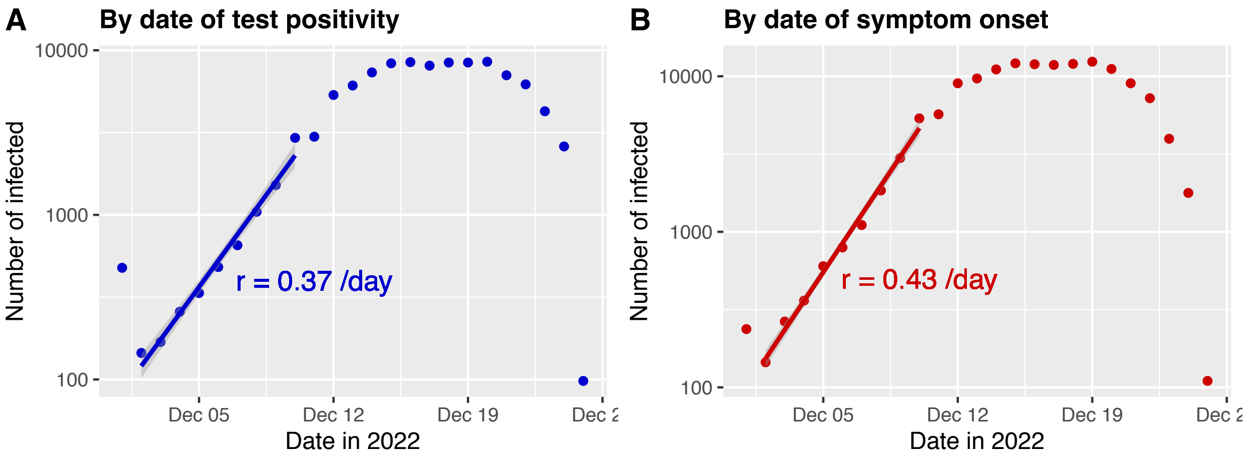


Figure S5: **Number of infected individuals by date of test positivity (A) or by date of symptom onset (B) in the Sichuan survey data.** The exponential growth rates, *r*, were estimated using data between Dec. 2 and 10, during which the number of infected people grew exponentially. Data on Dec. 1 was not used in estimation, because the numbers on the day likely include individuals who tested positive or had symptom onset before Dec. 1.

| period | increase parameter | median [95% CrI] |
|---|---|---|
| Oct 28 – Nov 11 | $\beta$ | 0.582 [0.546, 0.615] |
| | $r$ | 0.148 [0.129, 0.164] |
| | $R$ | 1.57 [1.49, 1.64] |
| Nov 11 – Dec 7 | $\beta$ | 0.618 [0.563, 0.681] |
| | $r$ | 0.165 [0.138, 0.195] |
| | $R$ | 1.65 [1.53, 1.79] |
| Dec 7 – Dec 26 | $\beta$ | 1.32 [1.09, 1.65] |
| | $r$ | 0.421 [0.351, 0.508] |
| | $R$ | 3.13 [2.66, 3.79] |

Table S1: **Estimated values for the transmission rate ($\beta$), intrinsic rate of increase ($r$) and reproductive number ($R$) during each modeled time period.** Note that the values of $r$ and $R$ reported for the period between Dec. 7 and Dec. 26 represent the growth rate and the reproductive number during the exponential growth of the outbreak.

| Province | Uninfected | Asymptomatic | Symptomatic | Recovered | Population Size (2021) |
|---|---|---|---|---|---|
| Anhui | 0.23 | 0.04 | 0.48 | 0.25 | 61,027,171 |
| Beijing | 0.19 | 0.01 | 0.24 | 0.56 | 21,893,095 |
| Chongqing | 0.20 | 0.03 | 0.31 | 0.46 | 32,054,159 |
| Fujian | 0.49 | 0.03 | 0.41 | 0.06 | 41,540,086 |
| Gansu | 0.22 | 0.03 | 0.39 | 0.36 | 25,019,831 |
| Guangdong | 0.30 | 0.02 | 0.45 | 0.23 | 126,012,510 |
| Guangxi | 0.31 | 0.04 | 0.46 | 0.19 | 50,126,804 |
| Guizhou | 0.26 | 0.03 | 0.56 | 0.15 | 38,562,148 |
| Hainan | 0.55 | 0.00 | 0.33 | 0.12 | 10,081,232 |
| Hebei | 0.19 | 0.01 | 0.26 | 0.53 | 74,610,235 |
| Heilongjiang | 0.24 | 0.02 | 0.53 | 0.21 | 31,850,088 |
| Henan | 0.19 | 0.02 | 0.36 | 0.43 | 99,365,519 |
| Hubei | 0.19 | 0.03 | 0.29 | 0.49 | 57,752,557 |
| Hunan | 0.20 | 0.04 | 0.51 | 0.25 | 66,444,864 |
| Inner Mongolia | 0.28 | 0.04 | 0.49 | 0.20 | 24,049,155 |
| Jiangsu | 0.27 | 0.02 | 0.53 | 0.18 | 84,748,016 |
| Jiangxi | 0.20 | 0.02 | 0.53 | 0.25 | 45,188,635 |
| Jilin | 0.29 | 0.01 | 0.39 | 0.31 | 24,073,453 |
| Liaoning | 0.24 | 0.02 | 0.45 | 0.29 | 42,591,407 |
| Ningxia | 0.22 | 0.04 | 0.52 | 0.22 | 7,202,654 |
| Qinghai | 0.25 | 0.01 | 0.42 | 0.32 | 5,923,957 |
| Shaanxi | 0.24 | 0.02 | 0.45 | 0.29 | 39,528,999 |
| Shandong | 0.30 | 0.02 | 0.47 | 0.21 | 101,527,453 |
| Shanghai | 0.36 | 0.02 | 0.44 | 0.18 | 24,870,895 |
| Shanxi | 0.28 | 0.02 | 0.44 | 0.27 | 34,915,616 |
| Sichuan | 0.18 | 0.02 | 0.33 | 0.47 | 83,674,866 |
| Tianjin | 0.23 | 0.02 | 0.36 | 0.39 | 13,866,009 |
| Xinjiang | 0.25 | 0.04 | 0.41 | 0.30 | 25,852,345 |
| Xizang | 0.26 | 0.03 | 0.45 | 0.26 | 3,648,100 |
| Yunnan | 0.35 | 0.03 | 0.34 | 0.29 | 47,209,277 |
| Zhejiang | 0.32 | 0.03 | 0.49 | 0.15 | 64,567,588 |

Table S2: **Dec. 26, 2022 prevalence survey data, reported by RenSheTong [12].** Population sizes were taken from the National Bureau of Statistics of China (`http://www.stats.gov.cn/english/PressRelease/202105/t20210510_1817188.html`).

| **Q: Have you been infected with COVID-19? (If no test conducted, please reply based on your judgement.)** | | |
|---|---|---|
| Options | Votes | Percentage |
| Not yet infected. | 12,671 | 26% |
| Infected without symptoms. | 1264 | 2% |
| Infected with symptoms. | 19,345 | 40% |
| Infected and recovered. | 14,617 | 30% |
| **Q: For symptomatic infections, how severe was it?** | | |
| Options | | Percentage |
| Severe. | | 35% |
| Relatively severe. | | 41.1% |
| I felt okay. | | 15.9% |
| Relatively mild. | | 6.9% |
| Mild. | | 1% |
| **Q: For recovered patients, how long did the symptoms last?** | | |
| Options | | Percentage |
| 1-2 days | | 10% |
| 3-4 days | | 22.5% |
| 5-6 days | | 28.2% |
| 7-8 days | | 26.8% |
| 9-10 days | | 8.1% |
| 11+ days | | 4.4% |

Table S3: **Questions, options, and response results in the Dec. 26 nation-wide survey, reported by RenSheTong [12].**
